# Evaluation of Changes in Near Visual Acuity and Required Near Addition Power Following Systemic Chemotherapy

**DOI:** 10.1101/2025.08.15.25333758

**Authors:** Azam Abdollahi, Morteza Tabatabaeefar, Saeed Rahmani, Haleh Kangari, Mohammad Ghassemi-Broumand

## Abstract

**Background:** Near visual acuity (NVA) typically declines with age and after cataract surgery, and it can be influenced by various factors, including diseases and medications. The impacts of systemic chemotherapy on visual functions, particularly accommodation and NVA, have not yet been thoroughly investigated.

**Objective:** The present study is the first to investigate the effects of systemic chemotherapy on NVA and the required near addition power.

**Methods:** A total of 130 eyes from 65 cancer patients [52.3% (n = 34) women, 47.7% (n = 31) men] were evaluated for NVA and required near addition power before and after systemic chemotherapy. Data analysis was performed using SPSS software.

**Results:** The mean age of the participants was 52.63 (13.35) years (age range = 18-80 years). Before chemotherapy, the mean addition power was 1.58 (0.95) D, the mean uncorrected NVA (UCNVA) was 0.50 (0.36) the logarithm of the minimum angle of resolution (logMAR), and the mean corrected NVA (CNVA) was 0.05 (0.24) logMAR. After chemotherapy, changes in addition power were statistically significant (P < 0.05), although changes in NVA were not statistically significant (P > 0.05). UCNVA remained stable in 43.1% of cases, CNVA in 86.9%, and addition power in 46.9%.

**Conclusion:** Based on the findings of this study, systemic chemotherapy does not cause an uncorrectable reduction in NVA. However, it can affect the required near addition power. Further research is recommended to evaluate long-term visual changes in patients undergoing chemotherapy.

## Introduction

For various reasons, the incidence and prevalence of cancer are rapidly rising worldwide (1). Chemotherapy, a widely used method in cancer treatment, can inhibit the growth of cancer cells(2); however, it simultaneously causes numerous side effects on healthy body tissues, including the visual system .(3).

Visual functions decline with age. Both near visual acuity (NVA) and accommodative ability decrease as individuals get older(4, 5). Difficulty with NVA is commonly reported by patients over 38 to 45 years of age, a condition known as presbyopia (6). Despite available corrective methods, uncorrected presbyopia remains the leading cause of NVA impairment globally, affecting over a billion people worldwide(7-10). Presbyopia significantly impacts both quality of life and visual function (13-11). While common factors, such as age and physiological changes in the lens and ciliary body, contribute to this process (15, 14, 6), the effects of chemotherapy on accommodative power and visual acuity have not yet been fully investigated (16). Various factors can influence accommodation and, consequently, NVA (18, 17, 11, 4). Some studies have indicated that chemotherapy may affect vision-related structures (20, 19, 3)? but conclusive evidence regarding the extent and nature of these effects is still lacking.

Therefore, this study was designed and conducted to investigate changes in NVA [corrected NVA (CNVA) and uncorrected NVA (UNVA)] and the required near addition power following systemic chemotherapy. The findings of this research can be beneficial in evaluating the visual side effects stemming from chemotherapy and in providing better strategies to preserve visual function and enhance the quality of life for cancer patients.

## Methods

In this observational and cross-sectional study, 130 eyes from 65 cancer patients [52.3% women (n = 34) and 47.7% men (n = 31)] were examined to assess changes in NVA following systemic chemotherapy. The variables of uncorrected NVA (UCNVA) and corrected NVA (CNVA), and the required near addition power, were evaluated both before and after the first cycle of systemic chemotherapy. The ethics committee of Shahid Beheshti University of Medical Sciences (Tehran, Iran) approved the study protocol (ethics code: IR. SBMU.RETECH. REC. 1397.1006).

### Patient Selection

Patients diagnosed with cancer, whose condition was confirmed by a radiation oncologist and were presenting for their first systemic chemotherapy treatment, were invited to participate in the study. Willing patients, after meeting the inclusion criteria, underwent NVA assessment both before and after systemic chemotherapy. Patients aged 18 years and older were evaluated, regardless of cancer type or treatment regimen. The inclusion criteria for the study comprised a cancer diagnosis, initiation of systemic chemotherapy as the primary elective treatment, and a good performance status [Eastern Cooperative Oncology Group (ECOG) PS: 0-1]. The exclusion criteria were defined as general patient debilitation, the addition of radiotherapy to the treatment regimen, patient unwillingness to undergo testing, and patient death. Medication administration intervals varied from once weekly to once monthly (weekly, bi-weekly, tri-weekly, or monthly), depending on the specific disease type and severity, and drug regimen.

### Procedure

NVA was assessed monocularly at 40 cm using a near Snellen chart, both with and without positive addition, and the results were recorded in the logarithm of the minimum angle of resolution (logMAR). To determine the near addition power, after full distance vision correction, NVA was measured monocularly and subjectively. The minimum positive power required for complete NVA correction was then determined based on the age-related accommodative range and recorded in D (21, 18). A comprehensive ophthalmic and visual evaluation was conducted for each patient. To avoid pharmacological interference, no medications (such as cycloplegics or mydriatics) were used. All tests were performed in a seated position.

### Statistical Analysis

The data analysis was performed using SPSS software. The Shapiro-Wilk test was employed to assess the normality of the variable distributions. The before and after chemotherapy variables were found to not be normally distributed (P < 0.001). Therefore, non-parametric tests, specifically the Mann-Whitney U test, Kruskal-Wallis test, and Wilcoxon signed-rank test, were employed for data analysis. All data analyses were conducted at a significance level of α = 0.05.

## Results

A total of 130 eyes from 65 cancer patients (mean age = 52.63 ± 13.35 years, age range = 18-80 years) were examined. Of these patients, 52.3% (n = 34) were women and 47.7% (n = 31) were men, with 80% (n = 52) being over 40 years old. The most frequent age group was 50-60 years (32.3%). The most common cancers observed were gastrointestinal (32.3%) and breast (13.8%) cancers. The most frequently used chemotherapy regimens were Carboplatin [CAR] + Paclitaxel (Taxol) [PTX] (31.6%) and Fluorouracil (5-FU) + Oxaliplatin (Eloxatin) [OXA] + Leucovorin (Folinic Acid) [FOLFOX] (14.6%). Approximately 61.4% of patients had no underlying diseases other than cancer. Among those with co-morbidities, 16.6% had hyperlipidemia, 13.6% had diabetes, 13.5% had hypertension, and 10.6% had hypothyroidism, all of which were reported to be under control and treatment. Notably, 76.47% (n = 26) of women and 83.87% (n = 26) of men were over 40 years old. NVA statistics (before and after chemotherapy) are presented in Table 1.

**Table 1.**
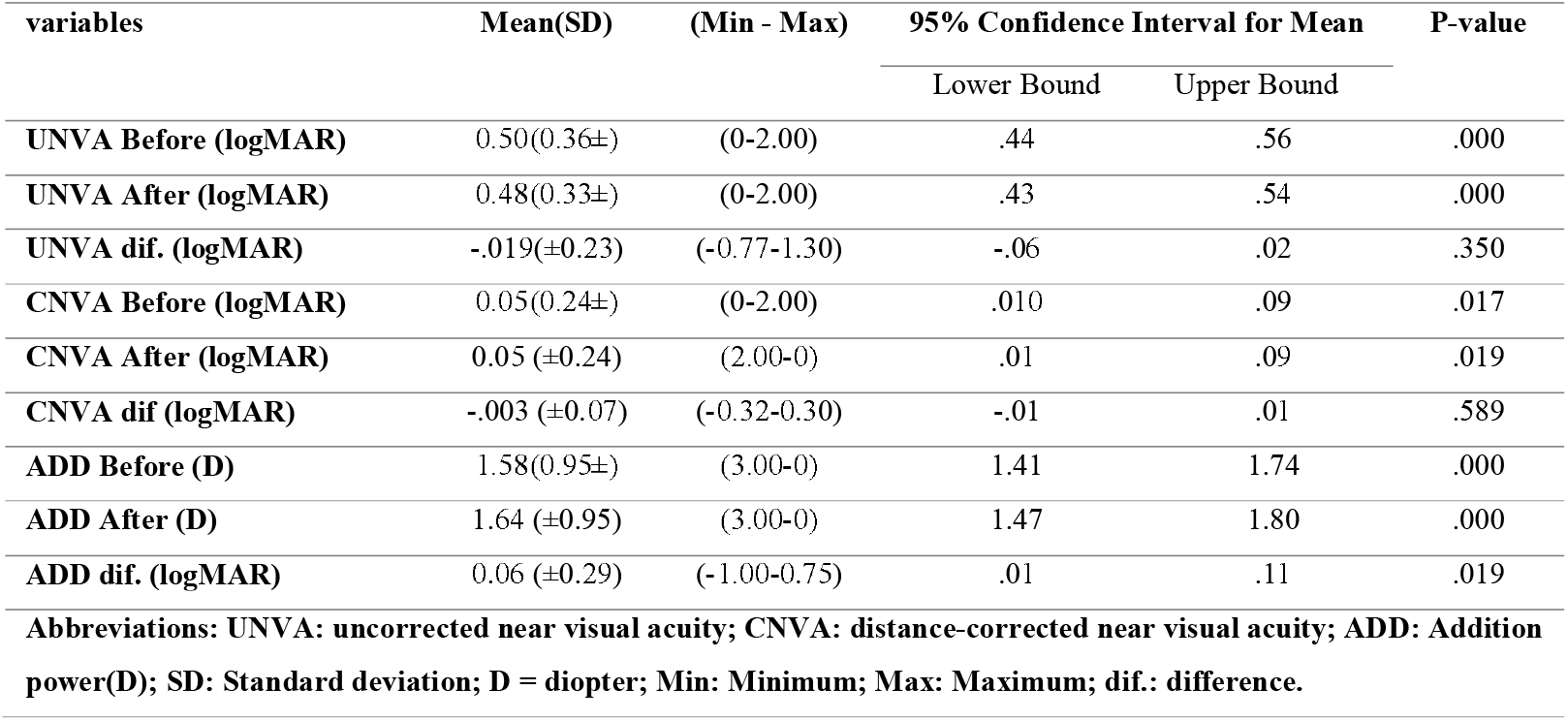
The statistical data (before and after chemotherapy) for near visual acuity and the required near addition power.

### Addition Power

Overall, 19.2% of eyes did not require positive addition power. Following chemotherapy, addition power increased in 33.8% of eyes and decreased in 19.2%. Incremental changes ranged from 0.25 to 0.75 D, while decremental changes ranged from -0.25 to -1.00 D. The most frequent decremental changes (13.1%) and the most frequent incremental changes (19.2%) were within ±0.25 D. Following chemotherapy, the frequency of the most common required addition, +2.00 D, increased from 16.9% to 17.7%. The lowest required addition, initially +0.25 D, shifted to +0.5 D after chemotherapy. The highest mean decrease in addition power was observed in the 70-80 age range, while the highest mean increase occurred in the 38-50 year old age range. The highest mean addition power was 2.77 D in the 70-80 year old age range. The most significant decrease in addition power was -1.00 D, and the most notable increase was 0.75 D, both observed in individuals over 40 years old. For those under 40, decremental and incremental changes in addition power were -0.25 D and +0.5 D, respectively. In the over-40 age group, decremental changes ranged from -0.25 D to - 1.00 D, while incremental changes in addition power varied from 0.25 D to 0.75 D. The most frequent (19.23%) incremental changes in addition power (0.25 D), followed by 0.5 D in 10.77%, both occurred in individuals over 40. Conversely, the most frequent decremental changes in addition power (−0.25 D) in 12.31%, succeeded by approximately -0.5 D in 5.39%, also occurred in individuals over 40 years old. Among individuals under 40, 3.85% experienced a decrease and 7.69% an increase in addition power. For those over 40, 23.08% exhibited a decrease and 40.39% an increase in addition power.

The addition power reduction observed in women ranged from -0.25 to -1.00 D, while in men, it ranged from -0.25 to -0.5 D. Conversely, the addition power increase in women ranged from 0.25 to 0.75 D, and in men, it ranged from 0.25 to 0.5 D. Overall, women exhibited a 14.71% decrease and a 42.65% increase in addition power, whereas men showed a 24.19% decrease and a 24.19% increase.

The highest mean increase in addition power was 0.50 D with PTX, while the highest mean decrease was -0.25 (±0.35) D in the Folinic Acid, 5-FU, and Irinotecan (FOLFIRI) regimen. Mean addition power changed by 0.10 (±0.26) D in the CAR + Taxol regimen and by -0.06 (±0.32) D in the FOLFOX regimen. Changes in addition power were incremental in the Doxorubicin (Adriamycin) + Cyclophosphamide [AC], Vincristine (Oncovin) [VIN], OXA, Gemcitabine + Cisplatin [GC], and Leucovorin + OXA + Avastin regimens. Additionally, the changes were decremental in the FOLFIRI, Bortezomib + Velcade, and Capecitabine [CAP] + OXA [CAPOX/XELOX] regimens. The changes were decremental and incremental in the Cisplatin, 5-FU, Cyclophosphamide + Doxorubicin (Adriamycin) + 5-FU [CAF], Leucovorin + Cytarabine + Methotrexate, CAR, and 5-FU + Leucovorin (Folinic Acid) [FL] regimens. In the CAR + Taxol regimen, addition power decreased by 14.71% and increased by 41.18%; in the FOLFOX regimen, it decreased by 33.33% and increased by 33.33%; and increased by 100% in the PTX regimen.

The greatest mean increase in addition power was observed in liver cancer [0.63 (±0.18) D], while the highest decrease occurred in laryngeal cancer [-0.25 (±0.20) D]. Addition power increased in brain, nasal, oral, bladder, and liver cancers. Conversely, it decreased in laryngeal, tongue, acute myeloid leukemia (AML), thyroid, and lung cancers.

In individuals without underlying diseases, the mean addition power was 1.38 (±1.01) D before chemotherapy, changing to 0.04 (±0.26) D following chemotherapy. The changes in addition power in individuals without underlying diseases exhibited an 18.42% decrease and a 31.58% increase.

### Uncorrected Near Visual Acuity (UCNVA)

The prevalence of individuals with complete UCNVA was 18.5%, which decreased to 16.9% following chemotherapy. Decremental changes (improvement) were observed in 31.5% of cases, while incremental changes (deterioration) occurred in 25.4%. Decremental changes (improvement) ranged from -0.02 to -0.77 logMAR, and incremental changes (deterioration) ranged from 0.02 to 1.3 logMAR. The most frequent improvement, at -0.3 logMAR, was observed in 6.9% of cases. The most frequent deterioration, at 0.17 logMAR, was observed in 8.5% of cases. No changes were recorded in individuals under 28 years of age. UCNVA changes in the 28-38 age range were 0.09 (±0.35) logMAR. The highest mean change was - 0.12 (±0.25) logMAR, with a minimum of -0.77 and a maximum of 0.17, in the 70-80 year old age group. The highest mean UCNVA before chemotherapy was 0.77 (±0.31) logMAR in the 70-80 age range. After chemotherapy, the highest mean UCNVA was 0.65 (±0.24) logMAR in the 70-80 year old age group and 0.65 (±0.15) logMAR in the 60-70 year old age group. The lowest UCNVA and the least changes were observed in individuals under 38 years of age. For those over 40 years old, improvements ranged from -0.02 to -0.77 logMAR, and deteriorations ranged from 0.02 to 0.47 logMAR. In individuals under 40 years old, improvements were within the -0.12 logMAR range, while deteriorations ranged from 0.28 to 1.3 logMAR. The most frequent improvement in UCNVA (6.92%) was approximately -0.3 logMAR, and the most frequent deterioration (8.46%) was approximately 0.17 logMAR among individuals over 40 years of age. UCNVA improved by 3.85% and deteriorated by 11.54% in individuals under 40 years, while in those over 40 years, it improved by 38.46% and deteriorated by 28.85%.

In UCNVA measurements, improvement-related changes were observed at 12.3% in men (ranging from -0.77 to -0.02 logMAR) and 19.3% in women (ranging from -0.60 to -0.02 logMAR). Conversely, deterioration-related changes were noted in 15.6% of women (ranging from 0.02 to 0.47 logMAR) and 10.1% of men (ranging from 0.02 to 1.3 logMAR). The frequency of UCNVA changes showed 36.77% improvement and 29.41% deterioration among women, and 25.81% improvement and 20.97% deterioration among men.

The highest mean increase in UCNVA (deterioration) was observed with the OXA regimen at 0.31 (±0.014) logMAR, followed by the PTX regimen with a mean of 0.28 logMAR. The highest mean improvement in UCNVA was found in an individual on both the AC regimen at -0.20 (±0.14) logMAR and the CAF regimen at -0.20 (±0.18) logMAR. The mean UCNVA changes were -0.06 (±0.33) logMAR for the CAR + Taxol regimen, with a minimum of -0.77 and a maximum of 1.3, and were -0.12 (±0.27) logMAR for the FOLFOX regimen, with a minimum of -0.6 and a maximum of 0.23. UCNVA changes showed improvement in the AC, FOLFIRI, CAF, Leucovorin + Cytarabine + Methotrexate, Leucovorin + OXA + Avastin, and Bortezomib + Velcade regimens. Moreover, deterioration was observed with the OXA, PTX, and VIN regimens. In the CAR + Taxol regimen, 44.12% showed improvement and 26.47% showed deterioration. For Cisplatin, 12.5% showed improvement and 37.5% showed deterioration. In the combination regimens of Cisplatin + 5-FU, FL, and XELOX, both decrease and increase were each 25%. In the GC regimen, UCNVA improved by 33.33% and deteriorated by 66.67%. Changes in UCNVA for the FOLFOX regimen exhibited 41.67% improvement and 16.67% deterioration.

The greatest mean increase in UCNVA (deterioration) was observed in individuals with rectal cancer as 0.31 (±0.01) logMAR. Conversely, the greatest mean decrease in UCNVA, (improvement) occurred in individuals with colon cancer as -0.11 (±0.29) logMAR). Changes in UCNVA showed improvement in AML and lymphoma cancers, while deterioration was noted in nasal, brain, laryngeal, and tongue cancers.

In individuals without underlying diseases, the mean baseline UCNVA was 0.46 (±0.38) logMAR, which changed to a mean of -0.01 (±0.23) logMAR. For such individuals, UCNVA improved in 26.32% and deteriorated in an equal percentage.

### Corrected Near Visual Acuity (CNVA)

Prior to chemotherapy, 89.2% of patients achieved a CNVA of 0 logMAR (full visual acuity), which decreased to 84.6% after chemotherapy. The most frequent changes observed were 0.02 logMAR in 5.4% of patients, while 86.9% experienced no change in CNVA. Decremental CNVA changes (improvement) ranged from -0.02 to -0.32 logMAR, and incremental changes (deterioration) ranged from 0.02 to 0.3 logMAR. Overall, CNVA improved in 5.4% of patients and deteriorated in 7.7%.

The highest mean CNVA before the intervention was 0.08 (±0.29) logMAR, with a minimum of zero and a maximum of 1.3, observed in the 50-60 age range. Conversely, the highest mean CNVA after the intervention was 0.09 (±0.20) logMAR, with a minimum of zero and a maximum of 0.70, found in the 70-80 age range. Regarding CNVA, the highest mean decrease was -0.02 (±0.06) logMAR, with a minimum of -0.3 and a maximum of 0.02, occurring in the 50-60 age range. Conversely, the highest mean increase was 0.02 (±0.13) logMAR, with a minimum of -0.28 and a maximum of 0.30, observed in the 70-80 age range. For individuals over 40 years of age, CNVA changes ranged from a deterioration of 0.02 to 0.3 logMAR and an improvement of -0.32 to -0.02 logMAR. Among individuals over 40, 6.73% showed improvement and 9.62% showed deterioration in CNVA. CNVA remained stable in individuals under 40 years of age.

The maximum improvement in CNVA was -0.32 logMAR in men, while the maximum deterioration was 0.3 logMAR in women. Regarding changes in CNVA, women exhibited a 4.41% improvement and 11.77% deterioration, whereas men exhibited a 6.45% improvement and 3.23% deterioration.

The highest mean CNVA deterioration was observed in individuals on a GC regimen, measuring 0.05 (±0.12) logMAR. Conversely, the greatest mean CNVA improvement occurred with the OXA regimen, at -0.05 (±0.07) logMAR. Mean changes in CNVA were - 0.01 (±0.10) logMAR in the CAR + Taxol regimen, -0.002 (±0.01) logMAR in the FOLFOX regimen, and -0.04 (±0.11) logMAR in the Cisplatin regimen. Among individuals receiving the CAR + Taxol regimen, 8.82% experienced improvement in CNVA, while 14.71% experienced deterioration. In the GC regimen, 33.33% of cases showed deterioration, with the remaining cases exhibiting stability. The FOLFOX regimen resulted in 9.09% improvement and 91.67% stability. For the OXA regimen, there was an equal distribution of 50% improvement and 50% stability. In the CAR + Taxol regimen, improvement-related changes ranged from -0.18 to -0.3 logMAR, while deterioration-related changes ranged from 0.18 to 0.3 logMAR. Both the GC and Cisplatin regimens demonstrated deteriorations.

The highest mean increase in CNVA (deterioration) was observed in an individual with bladder cancer [0.16 (±0.20) logMAR], while the highest mean decrease in CNVA (improvement) occurred in an individual with thyroid cancer [-0.16 (±0.23) logMAR]. After chemotherapy, CNVA worsened in patients with breast, bladder, intestinal, and liver cancers, while it improved in those with colon, rectal, and lung cancers.

In individuals without underlying diseases, the mean CNVA before chemotherapy was 0.04 (±0.24) logMAR, which changed by -0.01 (±0.05) logMAR. In such individuals, CNVA improved in 3.95% and deteriorated in 5.26% of cases.

Mean NVA (CNVA and UCNVA) and addition power based on age group and gender are presented in Table 2. The addition power before and after chemotherapy was significant based on age group and gender. Before and after the intervention, UCNVA was significant based on age group but not based on gender. Prior to the intervention, CNVA was not significant based on gender, but it became significant after the intervention based on gender (Table 2).

**Table 2.**
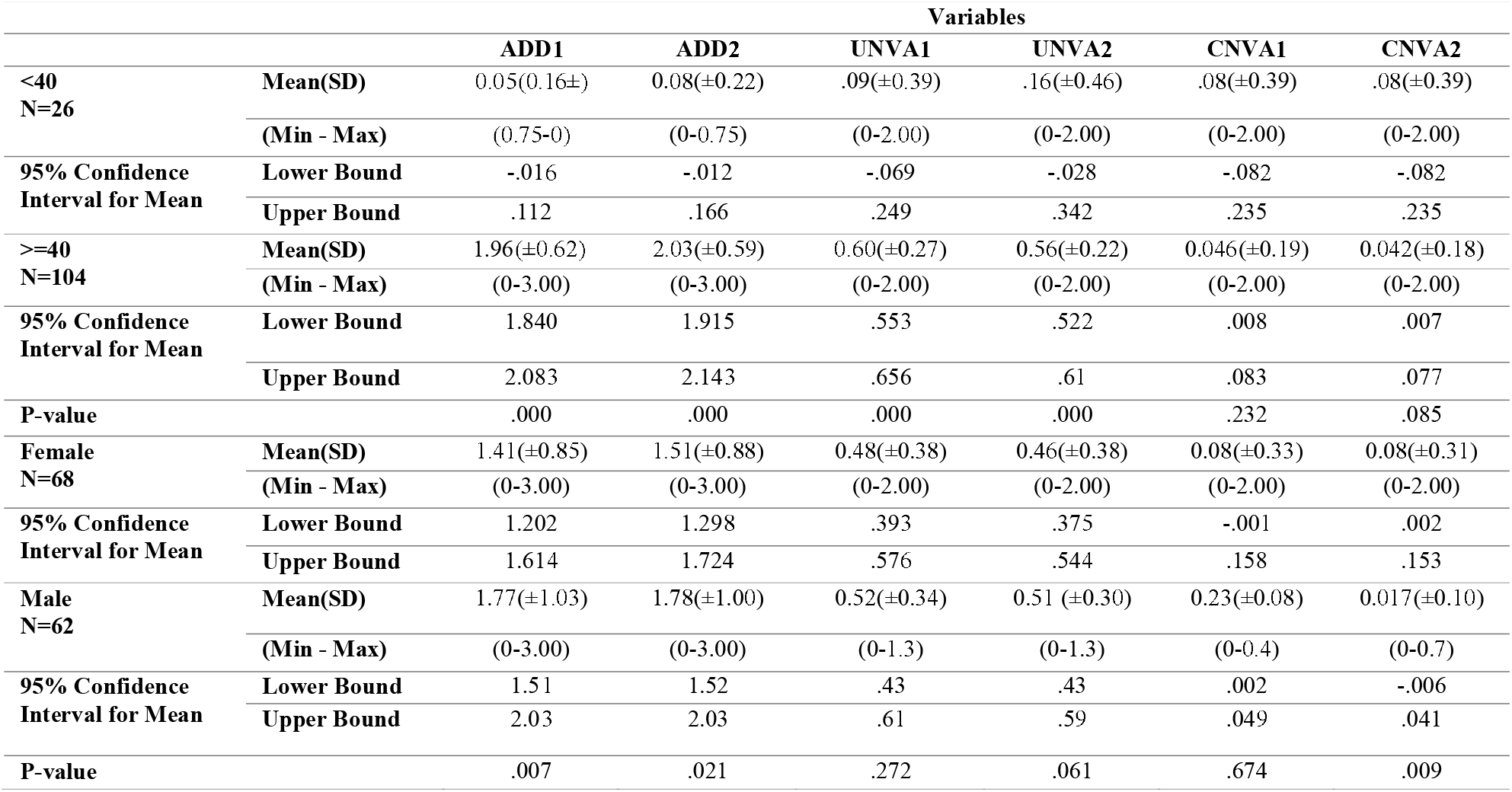
Mean (corrected and uncorrected) near visual acuity and addition power based on age group and gender.

Table 3 presents the mean changes in NVA (both CNVA and UCNVA) and addition power, stratified based on age group and gender. While the changes in addition power were not statistically significant across age groups, they were statistically significant based on gender.

**Table 3.**
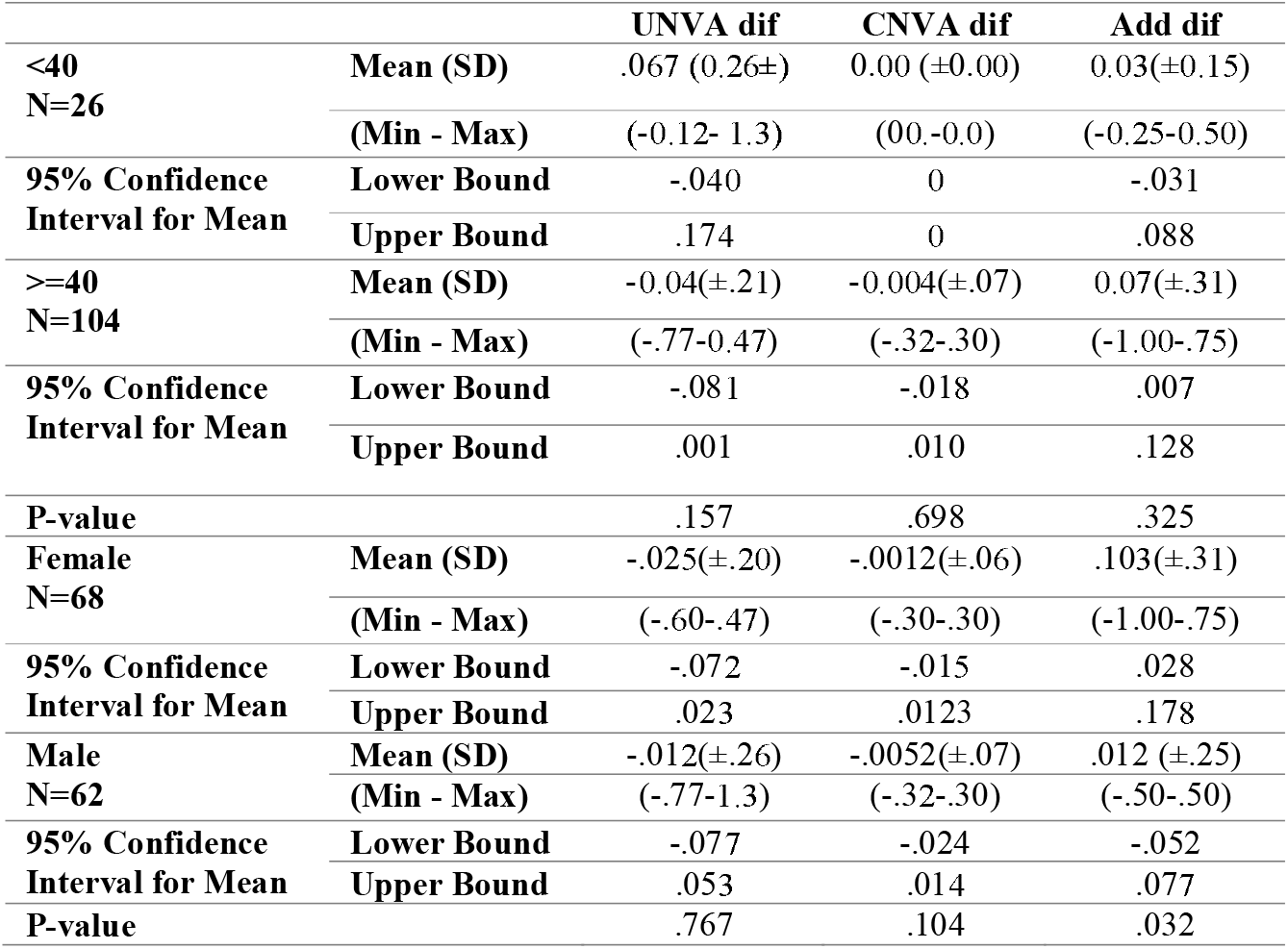
Mean changes in (corrected and uncorrected) near visual acuity and addition power based on age group and gender.

Figure 1 illustrates the amount of required addition power as a function of increasing age, demonstrating that addition power increases with age, particularly in individuals over 50 years old.

**Figure 1.**
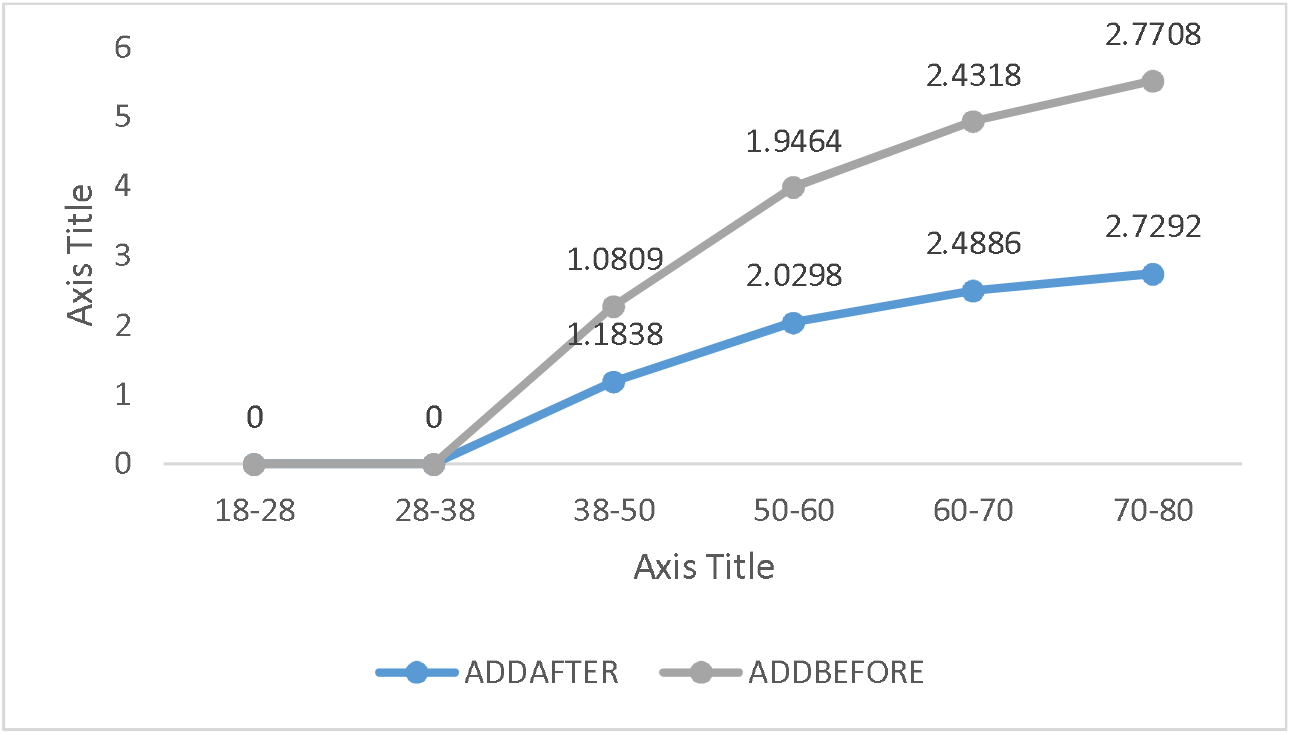
Changes in addition power with age

Figure 2 illustrates the frequency percentage of addition before and after chemotherapy, indicating an increased need for addition following chemotherapy.

**Figure 2.**
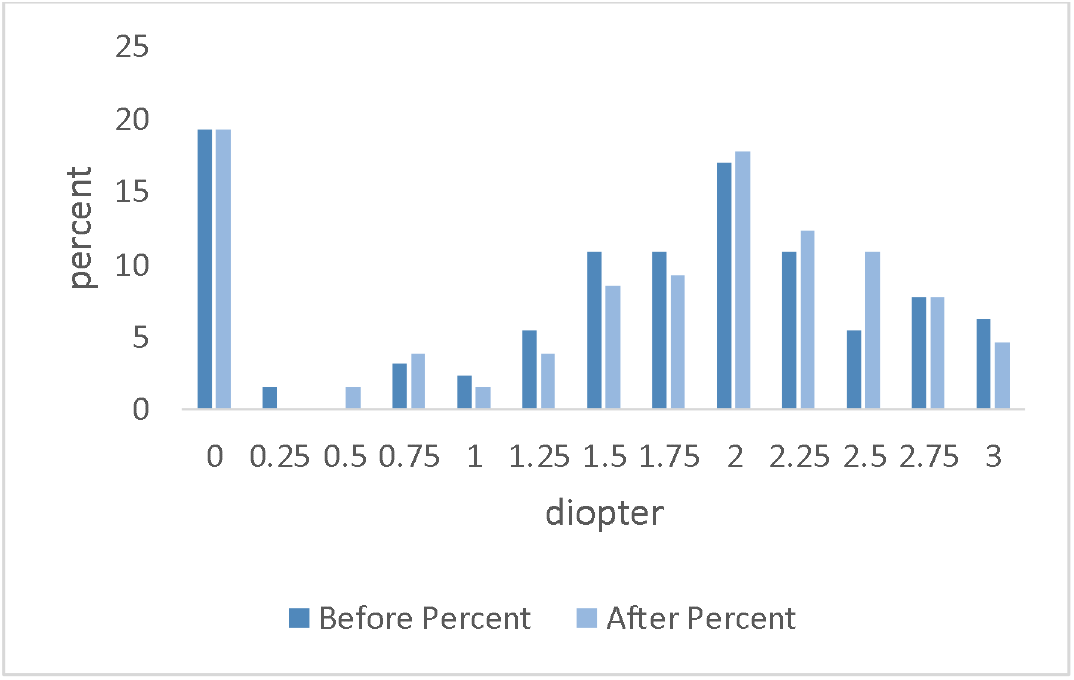
Frequency percentage of addition before and after chemotherapy

## Discussion

The mean addition power significantly increased after chemotherapy, indicating chemotherapy’s influence on factors affecting the required addition. The prevalence of incremental changes in addition power outweighed its decremental changes, which was expected given the age range of the majority of participants. Some observed incremental and decremental changes were beyond what could be solely attributed to age-related changes, potentially indicating drug side effects or disease progression. Following chemotherapy, the frequency of an addition of +2.50 D nearly doubled, a change unlikely to be solely due to aging. The most frequent increases and decreases were 0.25 D, which is a negligible issue, and can vary based on circumstances. The maximum required addition power was +2.00 D, which is typical considering the prevalence of the 50-60 year old age group and the amplitude of accommodation at these ages (18, 15, 4).

A minor, but not statistically significant, change in the mean UCNVA was observed following chemotherapy. Although some individuals experienced improvements and deteriorations of a few lines, research indicates that conventional chemotherapy does not significantly impact visual acuity (16). The changes in UCNVA after chemotherapy were relatively scattered, with both improvements and deteriorations noted, which may be attributable to the diversity of pharmacological regimens. Following chemotherapy, the prevalence of individuals with full vision decreased, which could be due to medication side effects or disease-related debilitation. Most individuals experienced changes that, given the short duration of the study, cannot be solely attributed to natural aging (5)(4).

The prevalence of individuals with complete CNVA significantly decreased following chemotherapy. The mean CNVA remained largely unchanged post-chemotherapy, and the frequency of minimum vision did not increase, suggesting that chemotherapy at conventional doses may be safe (Table 1). Overall, a substantial percentage of patients (86.9%) experienced no noticeable change in CNVA, although some cases showed a decrease or increase. Considering the predominant age range and corresponding adjustments, UCNVA and mean additions are also acceptable (18, 14, 4). The majority of individuals experienced no change in CNVA, indicating that most visual changes were correctable. Although improvements and deteriorations ranged from one letter to several lines, the most frequent changes were limited to a single letter, which is even considered normal in healthy individuals. However, with continued treatment, the extent of these changes may increase.

### Age and Changes

The mean addition power in individuals over 40 years of age showed a significant increase, which could indicate the influence of aging or medication (18, 15, 4). However, further investigation is needed to determine if the rate of these changes is directly correlated with age. The greatest increase in mean addition was observed in the 38-50 year old range, which is influenced by the progressive decline in amplitude of accommodation. (18, 4) The largest addition before and after the intervention was found in the 70-80 year old age range, which is acceptable due to the inability to accommodation within this age group. The most significant decrease in mean addition power was also observed in the 70-80 year old age range, which could be attributed to the onset of cataracts or medication side effects .(18, 5, 4).

The deterioration-related changes in UCNVA in individuals under 40 years old and its improvement-related changes in those over 40 could be attributable to the effects of chemotherapy. As anticipated, the worst pre-chemotherapy UCNVA was observed in the 70-80 year age range (18, 4). Contrary to age-related expectations, the highest frequency of improvement was found in the 38-60 year age range (18, 4).These discrepancies may stem from cancer type, specific pharmacological regimens, or individual patient characteristics.

Changes in CNVA were less pronounced than in UCNVA, which may be attributed to changes in refractive error or amplitude of accommodation. In individuals over 40 years of age, the prevalence of CNVA deterioration surpassed that of improvement, potentially indicating the onset of ocular diseases. The poorest post-chemotherapy corrected visual acuity was observed in the 70-80 year old age group, which is an expected finding given the age-related decline in visual functions and increased incidence of ocular diseases (5). The best NVA was noted in individuals under 38 years old, while the most significant changes were observed in the 50-60 year old age group, which is a normal finding. (18, 15, 4)

### Pharmacological Regimens and Changes

In this study, various pharmacological regimens had differential effects on addition power, necessitating a more detailed investigation into their pharmacological mechanisms. The patient treated with the PTX regimen demonstrated the highest increase, while the FOLFIRI regimen resulted in the greatest decrease. In the CAR + Taxol regimen, the mean changes in addition power were incremental. Medications can lead to changes in addition power by affecting the lens, accommodative muscles, or visual neural pathways (14, 16, 19). Further studies are required to determine whether certain chemotherapeutic agents exert a direct effect on ciliary muscle tone or alter lens structure, or if an indirect mechanism via the nervous system is involved. While limited, some chemotherapeutic drugs may induce retinal edema or changes in visual acuity (22, 20, 19, 3). Although some studies have reported an increase in hyperopia after chemotherapy, they have considered this effect to be negligible (16).

The highest mean UCNVA deterioration was observed in individuals receiving OXA followed by a PTX regimen. Notably, studies have reported visual acuity reduction with OXA (23, 20), and PTX has been shown to cause changes in the visual evoked potential .(24). There are also case reports of mild/moderate cystoid macular edema with PTX, subsequently leading to vision loss .(25, 22). Some studies indicate that visual changes following chemotherapy are primarily minor and clinically insignificant (16). .In CAR + Taxol and FOLFOX regimens, UCNVA improved, although deterioration was also observed. Rare case reports exist of optic nerve edema with hemorrhage and macular edema associated with CAR(19). However, in the current study, neither optic nerve nor macular edema occurred, suggesting that other contributing factors should be investigated.

UCNVA was slightly impaired with the Cisplatin + 5-FU regimen. However, this effect was not observed in CNVA, indicating that the changes were reversible. There are existing reports of retinal toxicity and vision loss associated with Cisplatin (26, 20). Vision deteriorated further with the OXA, PTX, and VIN regimens. Additionally, neuro-ophthalmic side effects have also been reported with VIN (27).

The mean CNVA improved in individuals undergoing the CAR + Taxol regimen; however, the frequency of deteriorations outweighed that of improvements. This observation aligns with previous studies where ocular deterioration was not unexpected (28, 19) Both Cisplatin and GC regimens exhibited deteriorations, consistent with prior reports of vision loss secondary to Cisplatin-induced retinopathy.(30, 29). Conversely, the FOLFOX and OXA regimens showed improvements, a finding that contradicts previous studies .(31). It is important to note that deterioration of vision was also observed in some individuals. This dual behavior in visual changes could stem from certain factors, such as cancer type and individual patient characteristics. Previous studies on the impact of chemotherapy on visual function have reported conflicting findings. Some research have demonstrated that specific drugs can lead to temporary or permanent visual changes(25, 20, 19), while others have reported only a limited effect.(16).

### Gender and Changes

The mean addition power was significantly lower in women than in men (P = 0.021) (Table 2). The frequency percentage, mean, and range of changes were higher in women than in men, which may be due to cancer type, pharmacological regimen, hormonal changes, or physical differences (33, 32). Moreover, a decrease in estrogen levels in women with increasing age culminates in changes in the lens (33).

The mean and frequency of UCNVA changes were higher in women than in men, although the range of changes was greater in men. Women have been reported to exhibit worse vision than age-matched men (35, 34). In women, 83.83% showed stability in CNVA, with a higher prevalence of deterioration-related changes compared to improvement-related changes. Conversely, 90.32% of men demonstrated stability, with improvement-related changes being more prevalent than deterioration-related changes. Further investigation is warranted to assess the extent and manner in which gender influences the accommodative system, visual function, and drug toxicity. Many gender-related differences in the eye have been attributed to the effects of sex hormones (36). Specific receptors for sex hormones, such as estrogen, progesterone, and androgen, have been identified in various structures of the anterior segment of the eye, including the lacrimal gland, cornea, lens, and ciliary body (39-37), with established roles of the ciliary body and lens in accommodation .(18, 15).

### Cancer Type and Changes

Following chemotherapy, the mean addition increased in individuals with brain and liver cancers, while it decreased in those with thyroid and laryngeal cancers. Although one study .(40) reported a higher prevalence of refractive errors in cancer patients compared to healthy individuals, there is no documented statistical data regarding presbyopia in this population. Further investigation is required to understand the reasons behind the varying deterioration-related and improvement-related changes in visual acuity observed in some patients, considering the location of their cancers. This investigation should separately examine the roles of pharmacological agents and the disease itself.

Significant limitations of this study included a high attrition rate due to patient withdrawal, disease progression, changes in treatment regimens, and patient mortality. Due to existing limitations, a detailed and comprehensive evaluation of various cancer types and pharmacological regimens was not feasible. Furthermore, given the specific conditions of the patients, investigating the precise mechanisms of visual changes using various imaging tests was not possible. Further research is required to elucidate the exact mechanisms underlying these changes.

The findings of this study open new research avenues in the fields of chemotherapy and visual function. These insights could be valuable in the visual management of patients undergoing chemotherapy. It is recommended that patients have regular ophthalmic examinations to monitor for any visual changes. Furthermore, the role of vision in assessing the side effects of treatment regimens should be emphasized. Future studies should focus on the long-term follow-up of patients and investigate the more sustained effects of chemotherapy on the eyes and vision.

Based on the findings of this study, while changes in NVA following systemic chemotherapy were not statistically significant, a notable increase in addition power was observed in some patients. Furthermore, visual changes were more pronounced in older age groups and among women. The effects of systemic chemotherapy on visual acuity, particularly in patients under treatment with specific drugs, such as OXA, PTX, and Cisplatin, warrant further investigation. Regular ophthalmic examinations are recommended for patients undergoing chemotherapy to monitor for potential visual changes. Future research should focus on the long-term assessment of visual changes in these patients.

## Data Availability

All data produced in the present work are contained in the manuscript

